# Detecting Shoulder Range of Motion Changes Following Breast Reconstruction Using a Novel Smartphone App

**DOI:** 10.64898/2025.12.22.25342831

**Authors:** Kayla Russell-Bertucci, Smarika Khadka, Kelly I. Nugent, Stephen M. Cain, Paige L. Myers, Adeyiza O. Momoh, Daniel L. Hertz, David B. Lipps

## Abstract

Breast reconstruction following mastectomy can restore body image and confer psychosocial benefits but can also result in lasting functional deficits in shoulder range of motion (SROM). Early identification of these deficits is crucial for guiding rehabilitation and improving long-term outcomes, but traditional assessment tools are often costly or impractical for routine clinical use. This longitudinal observational study examined the feasibility and validity of an app-based shoulder mobility assessment (MotionDetect) for detecting post-operative changes in SROM compared to traditional inertial measurement units (IMU)-derived measurements among breast cancer patients undergoing reconstruction. Twenty female participants undergoing bilateral mastectomy with immediate breast reconstruction performed at-home SROM assessments before surgery and once at 6–12 weeks post-operatively using both wearable IMUs and the MotionDetect iPhone application. Maximum shoulder abduction and flexion angles were recorded at each time point. Structured interviews gathered patient feedback on app usability. Statistical analyses assessed changes over time, correlation between measurement modalities, and repeatability. Significant reductions in abduction ROM after surgery were observed using both IMU and iPhone app assessments (both p < 0.031), with strong correlations between modalities (r > 0.80). Both approaches demonstrated excellent intra-class coefficients (ICC) repeatability (ICC > 0.89). Patient interviews indicated high feasibility and acceptability, with minor logistical challenges. Overall, this study indicates MotionDetect is a valid and feasible tool for remotely identifying post-operative SROM limitations in breast cancer patients, enabling early referral for rehabilitation to improve long-term quality of life.

## INTRODUCTION

Approximately 40% of the nearly 250,000 patients newly diagnosed with breast cancer in the U.S. each year undergo mastectomy^1, 2^. Women undergoing mastectomy will often opt to undergo breast reconstruction to restore the breast mound, which provides psychosocial benefits to patients^3^. However, mastectomy and reconstruction cause functional deficits in the shoulder, including 20-25% reductions in shoulder strength and shoulder range of motion (SROM) deficits of 15 – 25 degrees when compared to the preoperative state^4–10^. These deficits in SROM will ultimately impact the ability to perform everyday tasks. For example, female breast cancer survivors are almost three times more likely to stop working within a year of treatment than women in the general public, contributing to about $1 billion in annual economic loss^11, 12^. While this reduction in work capacity is multifactorial, breast cancer survivors who do not return to work report difficulties performing tasks such as reaching overhead^13^.

Earlier identification of SROM limitations during the postoperative period can benefit patients by ensuring they receive rehabilitation services sooner, before they are severely affected^14^. However, clinical assessments of SROM are rarely conducted during oncologic or surgical follow-ups due to their inconvenience, expense, and time requirements^15^. Additionally, goniometer- and inclinometer-based measures of SROM have highly variable reliability and are rarely completed independently by patients^16^. More accurate solutions for capturing shoulder kinematics include marker-based motion capture, which usually requires access to expensive equipment and a large laboratory setup. Movement sensors such as body-worn inertial measurement units (IMUs) can provide a robust examination of shoulder kinematics and have been used to track real-world arm use in clinical populations during everyday life^17–19^. While informative for research purposes, IMUs are still too expensive and inconvenient to use for clinical screening of postoperative deficits. Therefore, there is a need to develop new, low-cost tools for monitoring the shoulder health of patients after breast cancer treatment that could be performed with minimal to no clinical supervision.

A cost-effective solution for monitoring SROM after breast cancer surgery are smartphones. Smartphones are nearly ubiquitous, with >80% of adults owning one in the U.S.^20^. Mobile applications (apps) are an attractive approach for conveniently collecting toxicity data^21–23^. Apps can leverage biosensors within smartphones to collect objective data on movement disorders across many disease states^24, 25^. These app-based assessments could be a convenient and scalable strategy for managing chronic side effects both during and after cancer treatments. They could also be integrated into telehealth practices to provide more accessible, faster care without the need for an office visit. For example, mobile applications could help oncologists identify patients who experience post-mastectomy reductions in SROM and refer them to occupational therapy to improve SROM and long-term quality of life.

Therefore, the objective of this longitudinal observational study is to determine whether reductions in SROM can be identified via app-based assessment (MotionDetect). We further compare MotionDetect with IMU-derived assessments of SROM and gather patient feedback on their experiences using MotionDetect. We hypothesized that there would be decreases in shoulder abduction and flexion range of motion following surgery since previous studies reported arm mobility restrictions. We further hypothesized that the MotionDetect app would detect similar reductions in SROM after surgery as wearable IMUs and reveal a strong correlation between the two measurements. Overall, MotionDetect could be a convenient functional assessment tool used for earlier detection and intervention of SROM limitations in patients for whom clinical assessment and management would be appropriate, thus improving postoperative outcomes.

## MATERIALS AND METHODS

### Participants

Participants were recruited from the Division of Plastic Surgery at the University of Michigan who elected to undergo unilateral or bilateral breast reconstruction following mastectomy for breast cancer or prophylactic reasons. Female participants were included who were over the age of 18, with breast reconstruction performed immediately at the time of mastectomy with a two-stage implant or tissue based reconstruction. Participants needed access to an iPhone to participate in the study, as an alternative Android version of the application did not exist at study initiation. Participants were excluded if they had previous breast augmentation surgery or previous orthopedic or neurologic injury resulting in chronic (>1 year) reductions in arm function. Participants self-reported hand dominance. All participants completed registration and signed an informed consent form within the MotionDetect application. The study protocol was approved by the University of Michigan Institutional Review Board (HUM00227263).

Enrolled participants were mailed a kit containing three IMUs to be worn on their upper arms and thorax and were also emailed instructions to use the MotionDetect iPhone application through the MyDataHelps platform (CareEvolution, Ann Arbor, MI). Participants performed at-home SROM assessment before breast reconstruction surgery, and again 6-12 weeks after their surgery. SROM was simultaneously measured using APDM Opal IMUs (Clario, Portland, OR) (containing 3-axis accelerometer (±200 g), 3-axis gyroscope (±2000°/s), and 3-axis magnetometer (±8 Gauss)) and an Apple iPhone application (MotionDetect) (**Figure 1)**. Participants downloaded MyDataHelps from the Apple App Store to their personal devices, which required an iOS 15.0 or later operating system. Participants were given a unique invitation to complete the consent form, screener, and the MotionDetect app within the MyDataHelps platform. MotionDetect first provided video and verbal instructions for donning the IMU sensors and holding their iPhone for reliable comparisons, including holding the phone with the screen facing their palm, and keeping their elbow and wrist locked while moving their arm out to the side (abduction) or in front of them (flexion). Participants were then instructed to perform a calibration with both devices, where their arms were held straight out at shoulder height to identify 90 degrees of shoulder elevation. Then, participants performed 3 repetitions of maximum shoulder abduction and shoulder flexion, where the maximum value was extracted for analysis. Verbal instructions over the phone’s speaker were provided to let participants know when to start and stop each movement.

**Figure 1:**
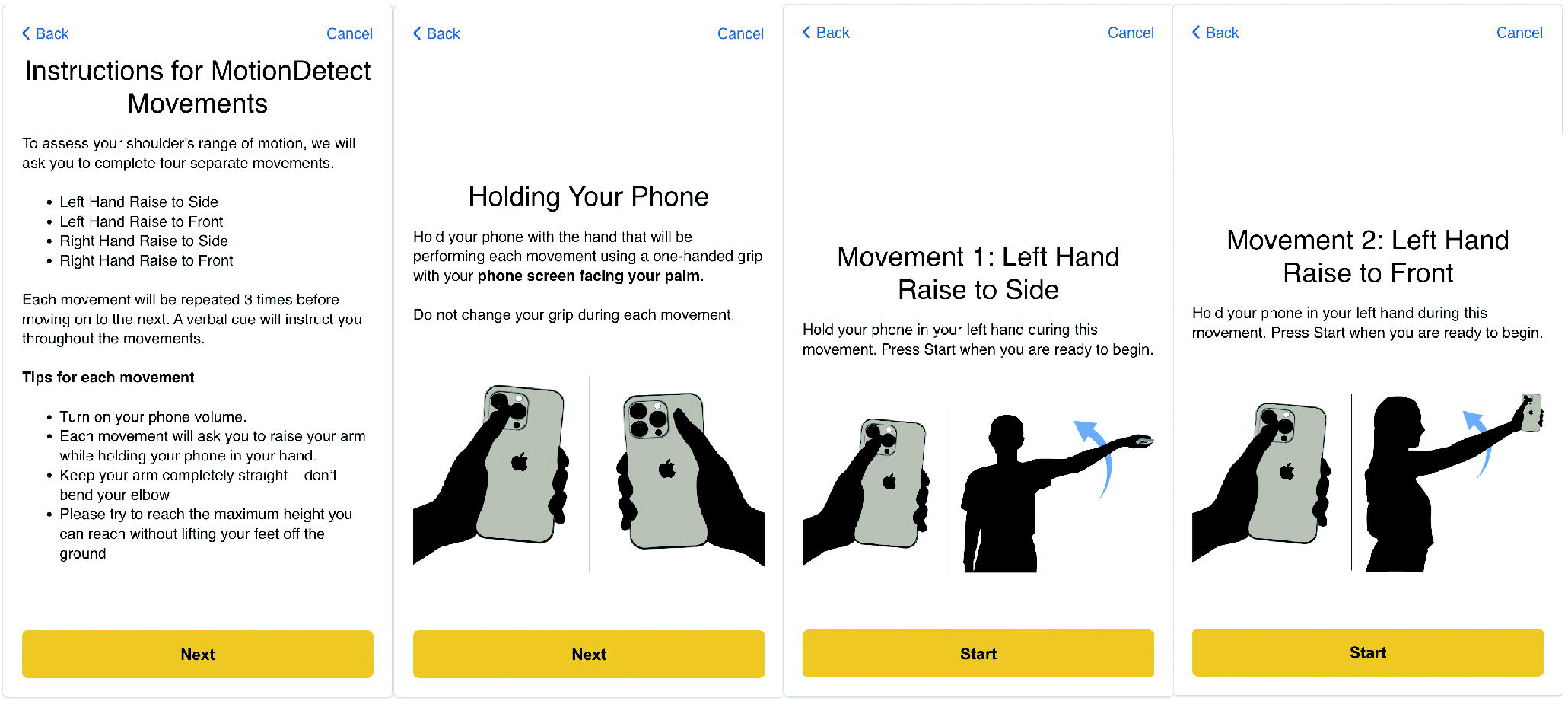
Screenshots from the MotionDetect iPhone app within the MyDataHelps platform depicting instructions for movements (first panel), instructions for holding the phone with the screen towards the palm (second panel), and instructions to complete arm movements to the side (third panel) or to the front (fourth panel)

### Structured Interviews

After completing the study, patients were invited to participate in an optional structured interview to discuss any issues they experienced while using the app. The structured interview addressed five key topics: (1) enrollment and app downloading, (2) ease of completing MotionDetect assessments, (3) timing and frequency of assessments, (4) total time required to complete assessments, and (5) interests in receiving information returned by the app (**Table 1**).

**Table 1:** Structured interview guide.

|  |  |
| --- | --- |
| **Enrollment and app downloading** | What challenges, if any, did you experience with downloading the MotionDetect app or enrolling in the study? |
| **Ease of completing mobile app assessments** | Which MotionDetect assessments were difficult or uncomfortable to complete? Why? |
| **Timing and frequency of assessments** | You completed MotionDetect assessments prior to and 4-8 weeks after surgery. Did you find this was too few or the right number of assessments? Was 4-8 weeks a good time for the post-surgery assessment or should it be sooner than 4 weeks or later than 8 weeks? |
| **Total time required to complete assessments** | How much time did it take you to complete the MotionDetect assessments each time? How much time do you think is reasonable to complete assessments each time? |
| **Interest in receiving information returned by the app** | If the app identified indications of reduced range of motion, would you be interested in having that information returned to you, shared with your doctor, shared with both you and the doctor, or not shared with either? |

### Data Analysis

The maximum shoulder elevation angle in flexion and abduction was measured from the IMUs and MotionDetect. A custom MATLAB (Mathworks, Natick, Massachusetts) code was written to calculate the humeral elevation using the acceleration and angular velocity data (frequency = 128 Hz) from the IMUs to estimate orientation. To calculate the humeral elevation angles, the functional calibration was used to align the inertial reference frame with the anatomical axes. Possible angles ranged from 0° (arm down at side) to 180° (arm raised above head). Further details regarding these IMU-derived calculations were published previously^19^.

MotionDetect used the Apple ResearchKit toolbox to measure changes in phone orientation (frequency = 100 Hz) relative to movements centered on the shoulder given the arm was fixed with the elbow fully-extended. Built-in accelerometers and gyroscopes within the iPhone were used to record a DeviceMotion file for each repetition. A custom MATLAB code was written to extract the phone orientation (in quaternions) and calculate the quaternion-derived range of motion (relative to the starting position for each repetition). The quaternions were converted to Euler angles using a ZYX sequence that converted the measurements from the standard iPhone coordinate system to a coordinate frame relative to the shoulder, with rotations in Y corresponding to shoulder elevation.

### Statistical Analysis

The study was powered to determine if the MotionDetect app could detect a large, clinically meaningful difference in SROM (dz = 0.8) between pre- and post-operative SROM as shown in other studies^7^. Assuming alpha = 0.05 and 80% power, we would need a minimum of 15 participants to detect a large, standardized effect size (dz = 0.8) with a two-tailed paired t-test.

To test our primary hypothesis that SROM would decrease following breast reconstruction surgery, a two-way repeated measures ANOVA model was utilized to examine the within-subjects factor of time and to control for arm dominance as a potential confounding factor. Separate models were performed for SROM measures acquired from the IMUs and from MotionDetect. Observed partial eta-squared (*η*^2^) effect sizes were also calculated, with *η*^2^ values > 0.14 considered a large effect size and *η*^2^ values between 0.08 and 0.14 considered a moderate effect size. All analyses were performed in SPSS 31 (IBM Corp).

The agreement between the pre to post-operative reductions in SROM detected with MotionDetect and the wearable IMUs was assessed using Lin’s Concordance Correlation Coefficient (CCC)^26^. The CCC was used to evaluate the degree to which the paired observations fell on the 45-degree line of identity. The CCC was further decomposed into the Pearson correlation coefficient (r), representing precision, and the bias correction factor (C_b_), representing accuracy. A C_b_ of 1 indicates no systematic bias. Separate correlations were run for the dominant and non-dominant arms.

Intra-class coefficients (ICC, one-way random model) were used to assess the repeatability of SROM measures obtained with IMUs or the MotionDetect app at baseline. Separate ICC values were obtained for flexion and abduction movements and for the dominant and non-dominant arms. An ICC exceeding 0.75 is considered excellent^27^.

## RESULTS

### Enrolled Patients

A total of 20 patients consented to the study and completed data collections at both time points. This included 14 women undergoing mastectomy with immediate two-stage prepectoral implants and six women undergoing mastectomy with immediate autologous deep inferior epigastric perforator DIEP flap (**Table 2**). All patients elected to undergo bilateral procedures. In addition, 5 women consented to the study, but their data were excluded because they did not complete the post-operative observation (N=4) or because their data were not fully uploaded to the cloud server due to wireless connectivity issues (N=1).

**Table 2:** Summary of patient demographics.

| Variable | Value |
| --- | --- |
| N | 20 |
| Sex (Female / Male) | 20 / 0 |
| Age (years) | 45.4 (9.2) |
| Height (m) | 1.7 (0.1) |
| Weight (kg) | 88.2 (22.1) |
| BMI (kg/m<sup>2</sup>) | 32.0 (7.3) |
| Dominant side treated | 8 / 20 |
| Prophylactic mastectomy (Yes / No) | 6 / 20 |
| Reconstruction Surgery (DIEP / PRE) | 6 DIEP; 14 PRE |
| Weeks post-surgery at follow-up | 10.1 (2.5) |

Abbreviations: DIEP - Deep Inferior Epigastric Perforator flap; PRE – prepectoral implant-based reconstruction

### Post-operative changes in SROM

A two-way repeated measures ANOVA found significant differences between pre- and post-operative abduction SROM, whether measured by iPhone application (F_1,19_ = 5.399, p = 0.031, □^2^ = 0.221) or wearable IMU (F_1,19_ = 2.798, p = 0.004, □^2^ = 0.354) (**Figure 2**). There was no significant effect of time observed for flexion SROM with IMU (F_1,19_ = 2.798, p = 0.054, □^2^ = 0.182) or iPhone measurements (F_1,19_ = 2.798, p = 0.111, □^2^ = 0.128). No significant interaction effects or main effects of handedness were observed. Overall, these results indicate that abduction SROM decreases following breast reconstruction.

**Figure 2:**
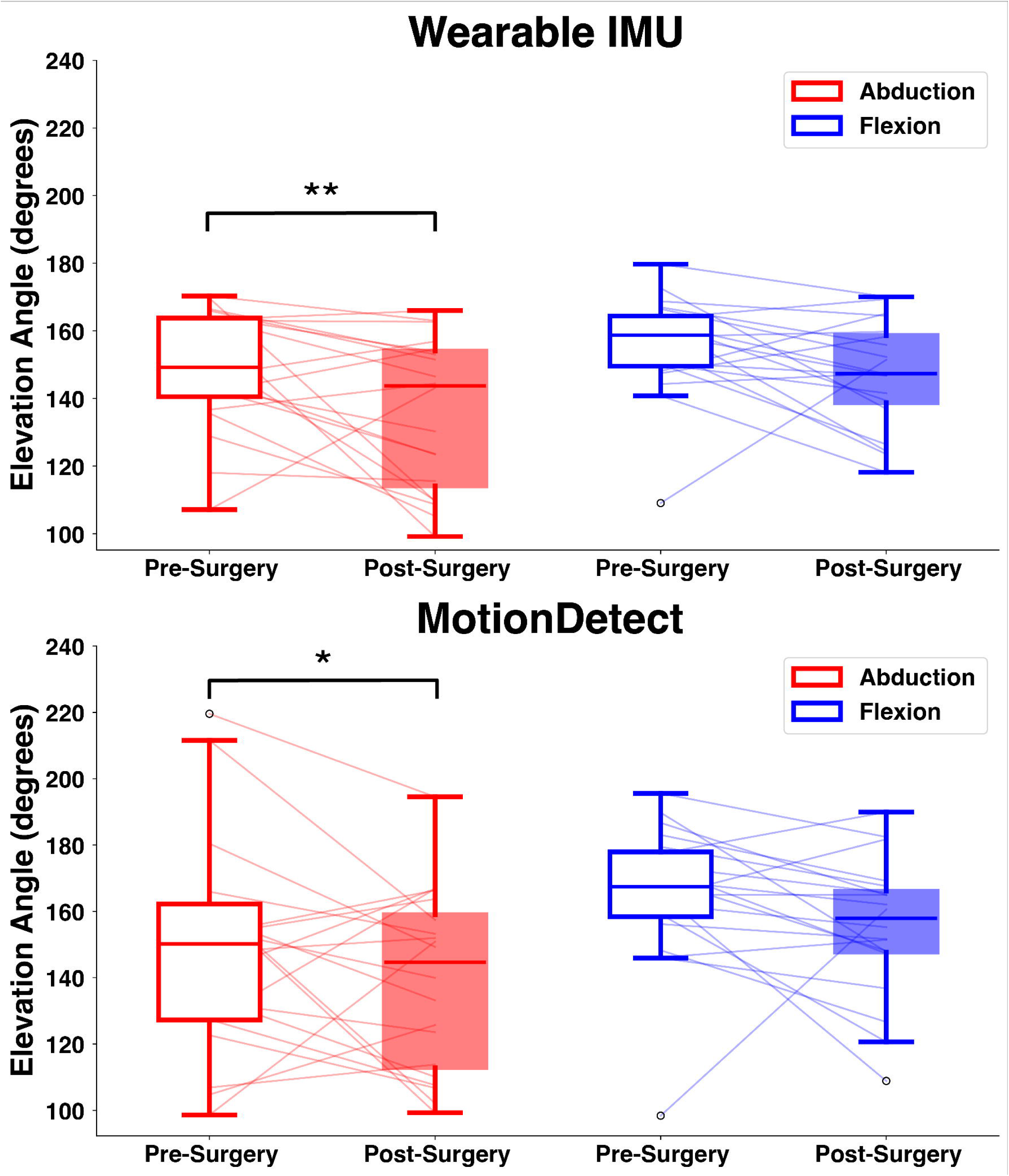
Dominant arm shoulder elevation angle recorded using wearable IMUs (top) and the MotionDetect iPhone app (bottom) as patients completed range of motion assessments in abduction (red) and flexion (blue) before and 8-12 weeks after mastectomy or immediate reconstruction. Significant differences are denoted with * if p < 0.05 and ** if p < 0.01.

### Comparison of MotionDetect and IMU performance

The analysis revealed substantial agreement in the pre- to post-operative change in flexion SROM measurements in the dominant arm with a CCC = 0.878 (95% CI: 0.750 – 0.942) with high precision (r = 0.909) and high accuracy (C_b_ = 0.910) between MotionDetect and wearable IMUs (**Figure 3**). The agreement for the non-dominant arm was moderate-to-substantial, with a CCC of 0.773 (95% CI: 0.549–0.893). The analysis revealed higher accuracy (C_b_ = 0.957) but lower precision (0.808) than the flexion SROM measures obtained from the dominant arm.

**Figure 3:**
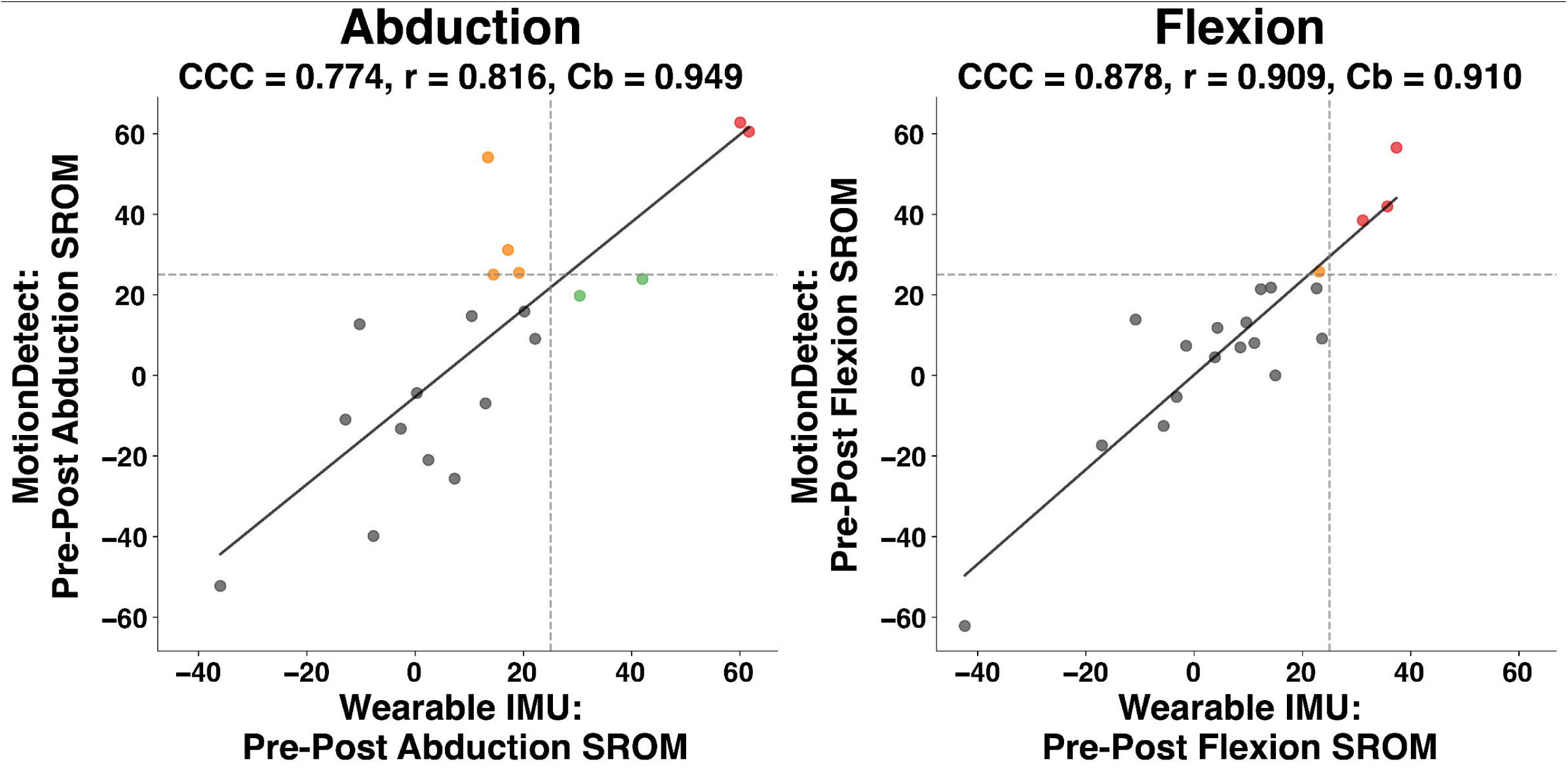
Comparison of the change in SROM between IMU-derived and MotionDetect-derived maximum shoulder abduction (left) and flexion (right). Changes are calculated as pre-operative minus post-operative. Clinically meaningful reductions in SROM (≥ 25°) are colored based on if MotionDetect (yellow), wearable IMU (green), or both (red) detected a deficit exceeding 25°.

Moderate-to-substantial agreement in the pre- to post-operative change in abduction SROM was observed for the dominant arm, with a CCC = 0.774 (95% CI: 0.551 – 0.894). The precision was less consistent (r = 0.816), but accuracy remained high (C_b_ = 0.949) between the two modalities. The agreement for the non-dominant arm was moderate (CCC = 0.671; 95% CI: 0.403–0.833). This lower concordance was characterized by a marked decrease in precision (r = 0.740) and a moderate decline in accuracy (C_b_ = 0.91).

### Repeatability

All measures showed excellent repeatability, with similar performance between SROM measures obtained with IMUs and with MotionDetect. For the wearable IMUs, ICC values for the non-dominant and dominant arms (respectively) during abduction were 0.98 and 0.96, and during flexion were 0.98 and 0.98. For the MotionDetect app, ICC values for the non-dominant and dominant arms (respectively) during abduction were 0.90 and 0.95, and during flexion were 0.89 and 0.95.

### Structured Interviews

Seven patients completed structured interviews about their experiences using the MotionDetect app. Overall, participants found the app and assessments easy to use, with few technical or logistical issues. Minor challenges were reported by three patients (43%), including difficulty locating the study within the app, limited cellular service, and the lack of Android availability. The arm-over-head assessment was reported as difficult or uncomfortable by two patients (29%) due to postoperative SROM limitations or surgical drains. Most patients (71%) felt that two assessments—pre-surgery and 6-12 weeks post-surgery—were appropriate, while three patients (43%) suggested adding a later assessment for those with delayed recovery. Completion time averaged 5–10 minutes and was considered reasonable by all participants (100%). All patients (100%) indicated interest in receiving their own results, and most (86%) also wanted their results shared with their physician. Despite minor technical or instructional issues, participants described MotionDetect assessments as feasible and acceptable within their recovery process.

## DISCUSSION

Impaired shoulder function after cancer treatment negatively impacts patients’ long-term quality of life^28^ and may contribute to the more sedentary lifestyles of breast cancer survivors following mastectomy^8^ or other cancer treatments^8, 29, 30^. Treatment-related adverse events affecting shoulder mobility are often undetected until patients are severely afflicted, supporting the need for clinical tools to identify at-risk patients sooner. This prospective study found that reductions in SROM in patients with breast cancer managed with mastectomy and immediate breast reconstruction can be identified via MotionDetect assessments from patients’ homes up to 12 weeks post-operatively. The MotionDetect app detected shoulder deficits with accuracy comparable to wearable IMUs placed on the upper body. Patient feedback regarding MotionDetect was mostly positive. These results support MotionDetect as a scalable, accessible clinical tool for evaluating SROM deficits.

SROM deficits in flexion and abduction are experienced by upwards of 2/3rds of breast cancer patients in the months and years after treatment^31–34^. While deficits immediately after surgery can be drastic (upwards of 59-80 degrees^34^), these reductions change over time and seem to settle to 15 – 25 degree reductions^4–8^. Clinically meaningful reductions in SROM >25 degrees are seen in ∼31% of patients at least three years post-mastectomy^7^. These effects may be more pronounced in breast cancer patients opting for post-mastectomy breast reconstruction^35^, leading to further reductions in shoulder strength and mobility^36, 37^. Our results indicate that remote patient monitoring of SROM is feasible with a mobile phone application and can detect clinically meaningful reductions up to 12 weeks post-operatively.

Convenient, cost-free digital health tools can scale the use of remote health monitoring in clinical care and research. >80% of Americans own a smartphone, including 79% of people older than 65 and 88% living in rural communities^38^, making mobile apps an attractive approach for developing digital health tools for convenient, scalable assessment of treatment-related adverse events^21, 39–45^. While the current study found similar SROM deficits after breast cancer surgery between wearable IMUs and MotionDetect, the cost and inconvenience of donning and wearing chest- and upper-arm IMUs limit their scalability. MotionDetect could be a more feasible and practical tool in clinical practice, despite some performance limitations relative to the wearable IMUs. Although long-term app persistence is suboptimal, short-term (< 3 months) persistence is high^46^, making this a feasible strategy for treatment-related adverse event assessment in the first three months after mastectomy. The current study shows promise for future inclusion in decentralized clinical trials and clinical care, particularly in rural environments, where patients face additional travel burdens and limited access to oncology rehabilitation specialists.

Smartphone apps have been developed and validated as an accessible measurement tool for range of motion and proprioception during lab testing. MotionDetect was remotely deployed, as breast cancer survivors tested their SROM before and after surgery while wearing IMUs without supervision. While other mobile applications have targeted SROM detection in breast cancer patients^47^, this study only examined healthy individuals in a laboratory setting, reaching to predetermined SROM angles (although in a larger variety of SROM directions than explored here). Therefore, ours is the first study to deploy an app to prospectively measure changes in SROM in breast cancer patients following treatment, while simultaneously validating these measures in patients’ homes. MotionDetect provided instructional videos in the app and verbal instructions, and participants did not require any clinical supervision while using the app. Despite the non-supervised remote deployment, patients achieved excellent repeatability with ICCs of flexion and abduction between 0.89 and 0.95, showing similar reliability to other validated smartphone accelerometer-based applications^47, 48^. Overall, MotionDetect demonstrated clinically meaningful reductions in two types of shoulder movement.

Scalable and accessible assessment tools, such as smartphone applications, have the potential to improve the effectiveness of rehabilitation strategies. The Centers for Medicare and Medicaid Services has developed procedure codes for remote therapeutic monitoring (CPT codes 98975-98981) that allow the use of digital data to assess treatment adherence and musculoskeletal pain. Further adoption of applications for remote therapeutic monitoring can assist case prioritization. For example, patients after mastectomy and breast reconstruction who are detected by MotionDetect to exceed SROM deficit thresholds can be referred earlier to therapeutic interventions than would be detectable with routine clinical follow-up. Clinically meaningful differences are typically interpreted when SROM deficits exceed 25 degrees for the gold-standard clinical assessment. Given the acknowledged variability in MotionDetect, a lower threshold between 15 to 20 degrees may be necessary for referral to for clinical assessment and then intervention if a SROM deficit is confirmed. Smartphone apps provide the advantage of more frequent data points due to short collection time. Increasing the frequency of these measurements may help detect when SROM changes occur following surgery and identify the optimal time frame for prescribing interventions when deficits are detected. Integrating SROM measurements from MotionDetect to existing toxicology smartphone apps^20^ may inform the development of a more comprehensive patient-reporting examination, enabling actionable clinical intervention and improved outcomes.

This study has several limitations. First, the current iteration of MotionDetect only supported Apple iPhones and did not support Android devices. Second, this was a feasibility study with a limited sample size of 20 evaluable patients recruited from two plastic surgeons at a single academic center. Third, the application had some issues with cellular service that led to problems in two cases with real-time data exports to a cloud server. The current iteration of the app did not store a local copy of the data, so it was not retrievable. Future iterations of the app should be better equipped to handle issues related to limited cellular service. Fourth, participants completed their follow up visits within a range of 6 weeks due to constraints with clearance to use their arms, completing the visit before their implant exchange, and overall availability. More standardized follow up timepoints may show different SROM changes following surgical intervention. Fifth, data analysis was limited to flexion and abduction of the shoulder, despite breast cancer survivors displaying decreases in internal and external rotation following similar treatments^9, 10^. Similarly, calculations were relative to gravity which cannot account for compensations occurring at the torso. Sixth, this study lacks direct comparison to clinical assessments of SROM measurements, which were not feasible as all patient interactions were performed remotely. Finally, future iterations of the application will allow for more comprehensive treatment-related adverse event tracking, including integration with NeuroDetect^22, 23^, an app-based assessment of chemotherapy-induced peripheral neuropathy.

## Conclusion

MotionDetect shows strong potential as a scalable, accessible, and convenient tool to measure SROM in oncologic or surgical settings. Measuring SROM following breast reconstruction should be a priority, as clinically meaningful deficits were observed in 40% and 20% of participants in abduction and flexion, respectively. Importantly, MotionDetect detected these deficits with strong correlations compared to wearable IMUs. However, MotionDetect should be tested on Android devices and deployed to a larger population prior to clinical implementation. Overall, this study indicates MotionDetect is a valid and feasible tool for remotely identifying post-operative SROM limitations in breast cancer patients, enabling early referral for rehabilitation to improve long-term quality of life.

## ACKNOWLEDGEMENTS

Financial support for this work was provided by American Cancer Society grants TLC-22-191-01 (DLH, DBL) and RSG-20-016-01-CCE (DBL). We also acknowledge the assistance of Anna Lamport with recruitment and data collection, and the support of CareEvolution in developing the mobile application.

## STATEMENTS AND DECLARATIONS

### Ethical considerations

This study was approved by the Institutional Review Board of the University of Michigan Medical School (HUM00227263) on February 6, 2023. This research was conducted ethically in accordance with the World Medical Association Declaration of Helsinki.

### Consent to participate

All participants provided written informed consent prior to enrollment in the study

### Consent for publication

Not applicable

### Data availability

The datasets generated during and/or analyzed during the current study are available from the corresponding author on request.

